# Costs and cost-effectiveness of Remifentanil Versus Fentanyl for analgo-sedation in Mechanically Ventilated Adult ICU Patients

**DOI:** 10.1101/2025.06.11.25329333

**Authors:** Sushma Ashwin, Lisa Gold, Arvind Rajamani, Ashwin Subramaniam

## Abstract

**Background:** It is common for intubated patients in intensive care units (ICU) to receive the opioids and sedatives infusions to relieve the pain and anxiety associated with invasive mechanical ventilation (IMV). A recent study demonstrated the feasibility of remifentanil. However, there is a notable lack of economic evaluations comparing these agents. This systematic review aimed to address this evidence gap.

**Methods:** In this systematic review, we searched Ovid MEDLINE, Ovid Embase, and the Cochrane Library for articles published between 1997 and 2023. We included full-text original English articles that reported on economic evaluation. We assessed the study quality using the 24-item Consolidated Health Economic Evaluation Reporting Standards (CHEERS) checklist.

**Results:** The search identified 169 abstracts; after removing duplicates, 103 were screened, and 8 full-text articles were assessed. Only two single-centre studies (n=285) met inclusion criteria as economic evaluations. The two studies varied significantly in methodology, including outcomes measured, cost components, time horizons, and perspectives. CHEERS checklist assessed the study qualities to be good (59%; Muellejans’) and very good (81%; Al’s). Although both studies demonstrated reduced IMV duration and ICU length of stay, the economic evaluation revealed conflicting results. While Al’s study found remifentanil may reduce costs, Muellejans’ study found similar total costs mainly due to higher drug prices of remifentanil.

**Conclusion:** This systematic review found only two heterogeneous studies on remifentanil’s economic impact in ICU, with inconsistent findings. The lack of robust evidence highlights the need for high-quality economic evaluations, given ICU care’s significant costs.

**PROSPERO:** CRD42022357343

## Introduction

In Australian intensive care units (ICU), over 50,000 critically ill patients require invasive mechanical ventilation (IMV) every year.^1^ It is common for intubated patients to receive the administration of both opioid analgesics and sedatives infusions to relieve the pain and anxiety associated with IMV (“analgo-sedation”).^1,2^ The ideal sedative would be cost effective, fast-acting, and free from adverse effects. However, among the currently used analgo-sedatives, there is no single agent that meets all these criteria. With prolonged use in critically ill patients, the commonly used opioids in Australian ICUs namely morphine and fentanyl exhibit potentially unfavourable pharmacokinetics such as long context-sensitive half-times resulting in drug accumulation, redistribution and residual sedation, particularly in patients with failing organs.^34^ This may lead to unpredictable effects such as prolonged deep sedation, immune suppression, hypotension,^5-7^ delayed weaning from IMV, potentially extending its duration and lengthening ICU stays.^8^

In contrast, remifentanil is an ultra-short acting opioid.^5^ It has a unique pharmacokinetic (**Table 1**) profile of a highly predictable onset and offset, organ-independent elimination and a short context-sensitive half-life.^9^ It elicits a deep analgesic state without tissue accumulation.^5,9^ A few randomised controlled trials (RCTs) have demonstrated that patients on remifentanil have resulted in reduced IMV duration, time to extubation after drug cessation, and ICU length of stay.^10-12^ A recent prospective, open-label pilot RCT comparing remifentanil to fentanyl in mechanically ventilated patients (>24 hours) found that remifentanil had a comparable or superior safety profile, with fewer complications, reduced delirium, and more ventilator-free days by day 28.^13^

**Table 1:** Pharmacokinetics of commonly used opioid medications.

| Medication | Onset (IV)<sup>1</sup> | Elimination half-life<sup>2</sup> | Context-sensitive half-life<sup>3</sup> | Route of administration<sup>4</sup> | Cost per unit<sup>5</sup> |
| --- | --- | --- | --- | --- | --- |
| Morphine | 5-10 min | 3-4 hours | N/A | IV, other methods | \$0.032 |
| Fentanyl | 1-2 min | 8-10 hours | 200 min (6-hour infusion); 300 min (12-hour infusion) | Subcutaneous, IM, IV, epidural, intrathecal, transdermal | \$4.936 |
| Hydromorphone | 5-15 min | 2-3 hours | N/A | IV, other methods | \$0.434 |
| Remifentanyl | 1-3 min | 3-10 min | 3-4 min | IV, intranasal | \$11.28 |
| Sufentanil | 5-10 min | 6-9 hours | 20-60 min | IV, other techniques including epidural, intrathecal, transdermal, and intranasal | \$5.27 |
| 1. The time it takes for a drug to start working after intravenous administration.<br>2. The time required for the drug's concentration in the bloodstream to decrease by half.<br>3. The time for a drug's concentration to halve after stopping a prolonged infusion, considering its distribution and redistribution in the body.<br>4. The method by which a drug is introduced into the body, such as orally, intravenously, or intramuscularly.<br>5. The cost of drug per mg in US dollars.<br>Abbreviations: IV – intravenous, IM – intramuscular, min – minutes, N/A – not applicable |  |  |  |  |  |

ICUs face ongoing pressure to deliver high-value care within limited resources. The COVID-19 pandemic has further highlighted the need to optimise ICU throughput and resource utilisation.^14,15^ Given the numerous priorities in healthcare delivery and the limited resources available for investment, it is crucial for decision-makers to assess the most effective options to optimise health outcomes. Since resource intensive treatments such as IMV could increase the daily costs by an additional 20-26%,^16,17^ decreasing the duration of IMV, and therefore ICU length of stay, may lead to clinical and economic benefits.^18^ Analgo-sedation strategies that enable faster weaning from IMV, such as using short-acting agents like remifentanil, may offer both clinical and economic advantages.^11,13^ While remifentanil shows promise in reducing complications and ICU length of stay compared to fentanyl, its higher acquisition cost raises questions about overall cost-effectiveness. The economic implications of choosing remifentanil over fentanyl are significant, particularly considering the high costs associated with ICU care, which can exceed A$4000 per patient bed-day.^19^ Will approximately 10% of these opioid costs have been attributed to pharmaceuticals,^20^ this has implications for healthcare policy and resource allocation, especially in a resource-limited system.^21^ Despite recognised clinical differences, there is a notable lack of economic evaluations comparing these agents. This systematic review aimed to address this evidence gap.

## Methods

This systematic review’s protocol was registered with PROSPERO (CRD 42022357343) before completing the search strategy development. The methodology of the review followed the guidelines outlined in the Preferred Reporting Items for Systematic Reviews and Meta-Analyses (PRISMA) statement.^22^

### Literature Search Strategy

A comprehensive search was carried out across key electronic databases, including Ovid MEDLINE, Ovid Embase, and the Cochrane Library. The search was limited to studies published after 1997, reflecting the year remifentanil was patented and its subsequent adoption in clinical practice. The search strategy was developed with support from a Deakin University librarian. The search was performed to capture the most recent studies and data available from 1997 to 2023. Search terms included a mix of controlled vocabulary (such as medical subject headings and Emtree) and free-text expressions related to remifentanil and fentanyl, together with terms associated with economic analyses like “cost”, as well as terms related to IMV and ICU (**Supplementary Table 1**). Additionally, the bibliographies of selected articles were reviewed to identify any further studies of interest.

### Study Selection

Screening was conducted by two reviewers (SA and AS) independently, with any differences resolved through consultation with a third reviewer (AR). The process involved two phases: initially, titles and abstracts were reviewed to filter out articles that did not meet the predefined criteria. The objective of the selection criteria, outlined in **Table 2** was to identify studies that were relevant to the study’s focus.

**Table 2:** Selection criteria for the systematic literature review.

| Inclusion criteria | Exclusion criteria |
| --- | --- |
| - Economic evaluations that were trial-based (deriving clinical and resource data from a single study) and model-based (incorporating data from various sources).<br>- Full economic evaluations including cost-effectiveness analyses (CEA, including Cost-Consequence Analysis (CCA) and Cost-Minimisation Analysis (CMA), Cost-Utility Analysis (CUA) and Cost-Benefit Analysis (CBA).<br>- Partial economic evaluation that performed cost analyses and reported outcomes in monetary units.<br>- Economic analyses which focused solely on costs and resources used, or which did not entail a comparator, were excluded.<br>- Published in the English language. | - Available in abstract form only (for example conference abstracts)<br>- Reviews of existing economic evaluations that did not present new data.<br>- Patient population not in ICU<br>- Results were not available separately for the ICU subgroup.<br>- Patient group - pregnant women, neonates or children or a mixed cohort where results were not available for the adult subgroup.<br>- Studies where remifentanyl/fentanyl were part of a variety of interventions being compared and could not be differentiated in terms of effects and cost of intervention. |

Subsequently, the full texts of the articles deemed to have been potentially eligible were thoroughly reviewed. Studies were restricted to those published in English. An economic evaluation was identified as being a comparison of different interventions regarding their costs (use of resources) and outcomes (effects, results).^21^ All studies that presented an economic analysis of analgo-sedation for adult patients undergoing IMV in the ICU were included. The reasons for excluding articles at this stage were systematically recorded.

### Data Extraction

The task of data extraction was carried out by two independent reviewers (SA and AS) using the Covidence platform (Veritas Health Innovation, based in Melbourne, Australia) and structured data extraction templates. Information gathered included specifics of the study (such as author, publication year and country of origin), details on the intervention and its comparators, and the methodology behind the economic evaluation (including aspects like the time horizon, currency used, discount rates, perspective of the study, type of economic evaluation, sources of data and any performed sensitivity analyses). The outcomes of interest focused on the cost-effectiveness of the interventions, determined through measures such as the incremental cost-effectiveness ratio (ICER), which could include costs per outcome achieved, costs per quality-adjusted life year (QALY), and costs per life year gained, among others. The outcomes of interest also included the likelihood of an intervention being cost-effective, the total costs involved, and the associated health outcomes, which included both mortality and QALY metrics. Interventions were categorised as having been “dominant” if they had produced lower costs and higher effectiveness relative to their comparators or were considered “dominated” if they had led to higher costs and reduced effectiveness.

### Data Synthesis

The studies were categorised based on the interventions that they had assessed. A qualitative analysis of the variability across trials was conducted to assess the feasibility of conducting a quantitative synthesis of the research findings. The primary outcome was cost-effectiveness for remifentanil when compared to a conventional opioid agent (fentanyl or morphine).

### Quality Assessment in Individual Studies

The assessment of study quality utilised the 24-item Consolidated Health Economic Evaluation Reporting Standards (CHEERS) checklist^23^ which was applied independently by two reviewers (SA and LG). Any disagreement between the two reviewers regarding the weightage was resolved through discussion. Each study received a score based on these 24 standards, with a full point awarded for each criterion satisfactorily met by the study. In line with the method used by Hope et al.,^24^ a criterion that was partially met resulted in a half-point award. For studies reported across multiple publications, all the relevant documents were reviewed during the quality assessment and the highest score obtained for any specific CHEERS item across these publications was recorded. The final quality score for each study was then expressed as a percentage. Based on their overall scores, studies achieving ≥85% were classified as exhibiting excellent reporting quality. Those with scores ranging between 70% and 85% were deemed to be of very good quality; those with scores between 55% and 70% were considered good quality; and studies that scored below 55% were considered of poor quality.^24^

## Results

The initial search yielded 169 abstracts. After removing duplicates, 103 unique abstracts remained for screening. Of the eight full-text articles assessed for eligibility, only two single-centre studies (n=285) met the inclusion criteria as health economic evaluations (**Table 3**). Results of the search strategy are illustrated in **Figure 1**.

**Table 3:**
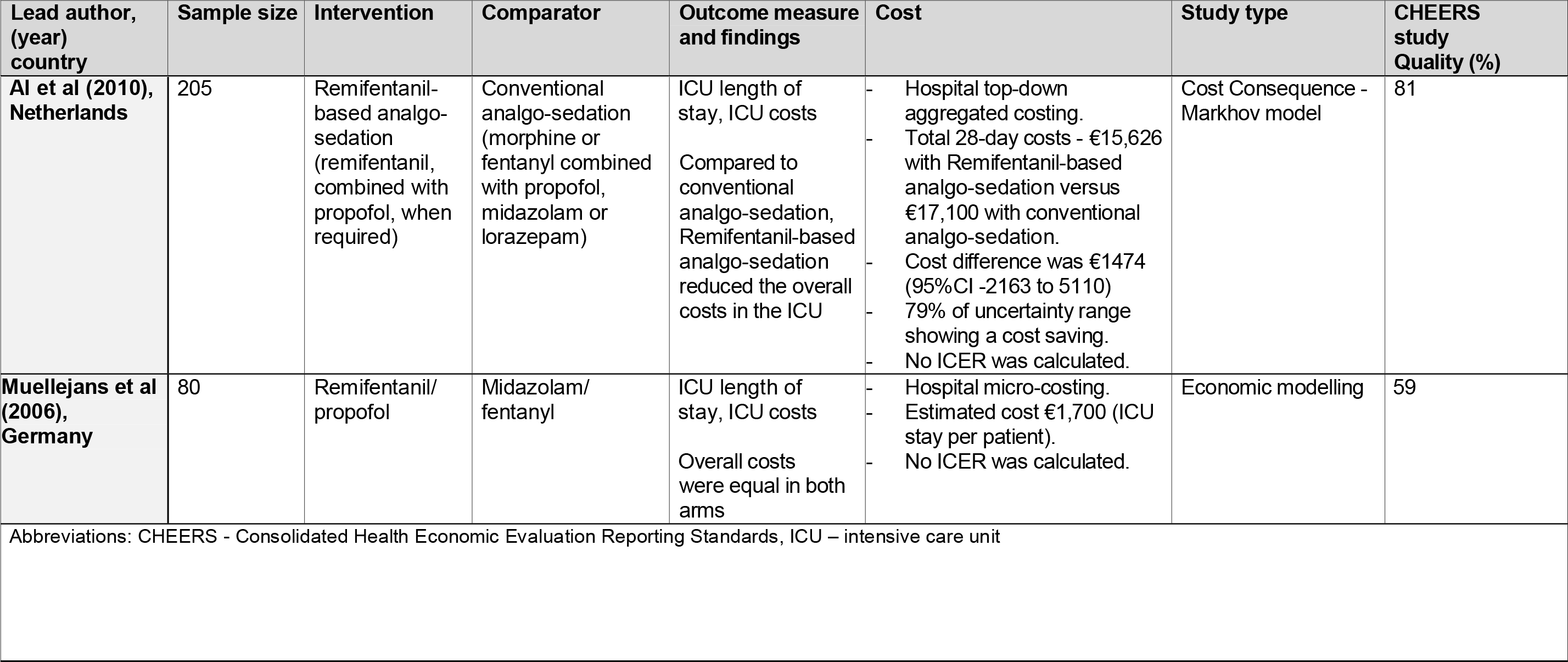
Characteristics of studies describing cost-effectiveness of remifentanil versus fentanyl in ICU patients.

**Figure 1:**
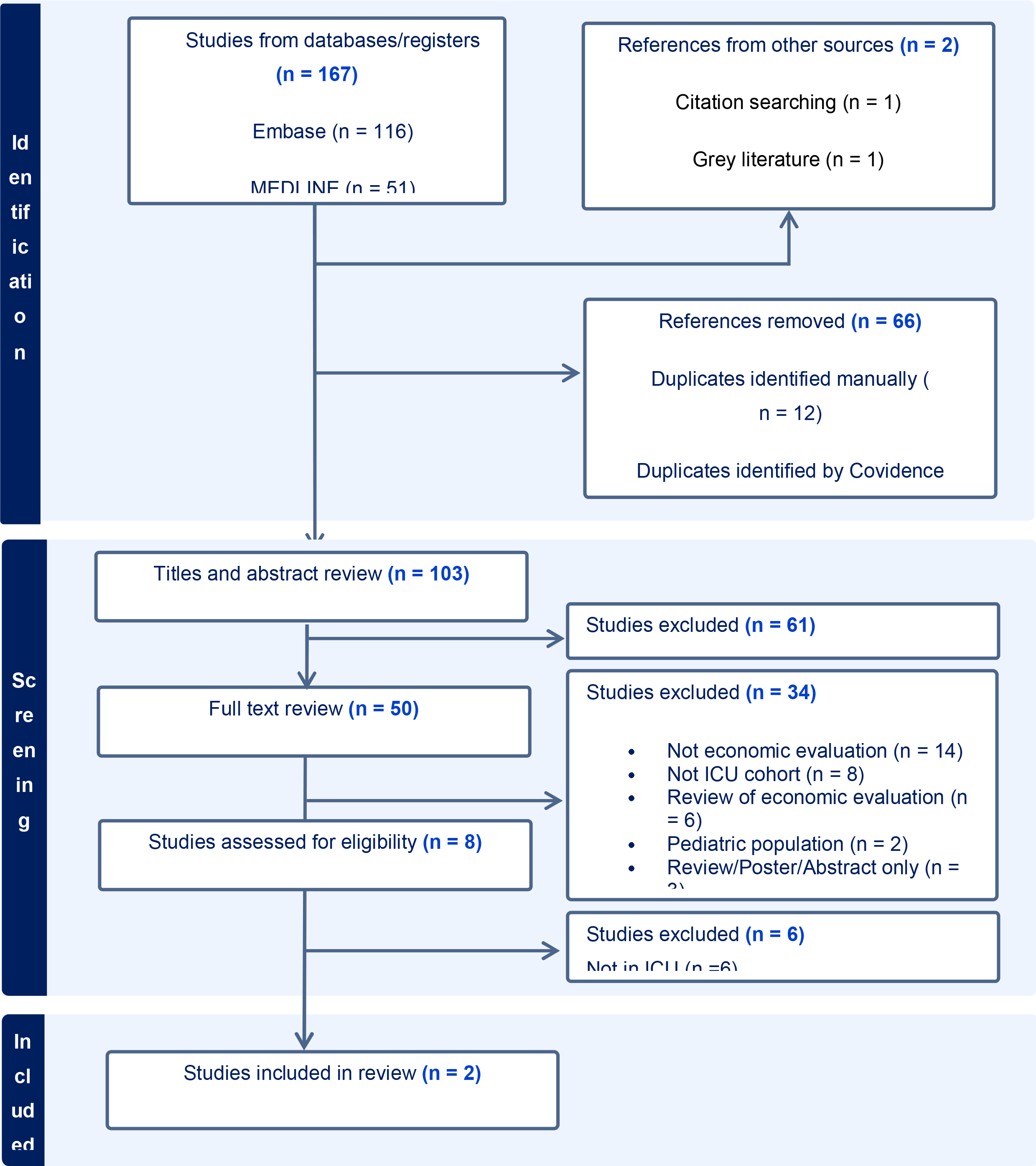
PRISMA Flow diagram to demonstrate study inclusions.

The Dutch study by Al et al.^11^ conducted a cost-consequence analysis comparing ICU length of stay between remifentanil-based and conventional analgo-sedation. It used data from the UltiSAFE trial^11^ and a separate micro-costing study,^17^ focusing on ICU patients requiring 2-3 days of IMV, from a hospital perspective using 2006 cost data. The analysis used a Markov model to estimate costs and outcomes beyond the 10-day remifentanil treatment studied clinically. It suggested that remifentanil-based analgo-sedation may reduce ICU length of stay and generate cost savings, despite a 79% uncertainty. Additionally, the study did not measure morbidity or mortality outcomes; it assumed an equal survival/mortality rate between groups. This assumption was based on the absence of differences observed in the clinical study, even though mortality had not been an outcome in the economic evaluation, which had focused solely on costs.

The German study by Muellejans et al.^12^ was a cost analysis that used modelling to explore deviations from protocol, particularly the high remifentanil dosage and its cost implications. This study did not directly compare the differences in costs to the differences in length of stay. Although the study perspective was not explicitly stated, it likely reflected a healthcare or hospital resource use perspective. The remifentanil/propofol regimen had significantly shorter weaning (2.2 vs. 5.7 hours, p<0.05) and extubation times (20.7 vs. 24.2 hours; p<0.05) when compared to the midazolam/fentanyl regimen. Despite higher remifentanil drug costs, total costs were similar. Sensitivity analysis supported these findings, and modelling suggested potential cost savings with adjusted remifentanil dosing in real-world practice.

### Quality Assessment

The CHEERS assessment of cost-effectiveness analysis is summarised in **Table 4**. The reporting quality of the two studies varied significantly: Al et al.’s^11^ was rated as very good quality study (81%), whereas Muellejans et al.’s^12^ study was only rated as good quality (59%).

**Table 4:**
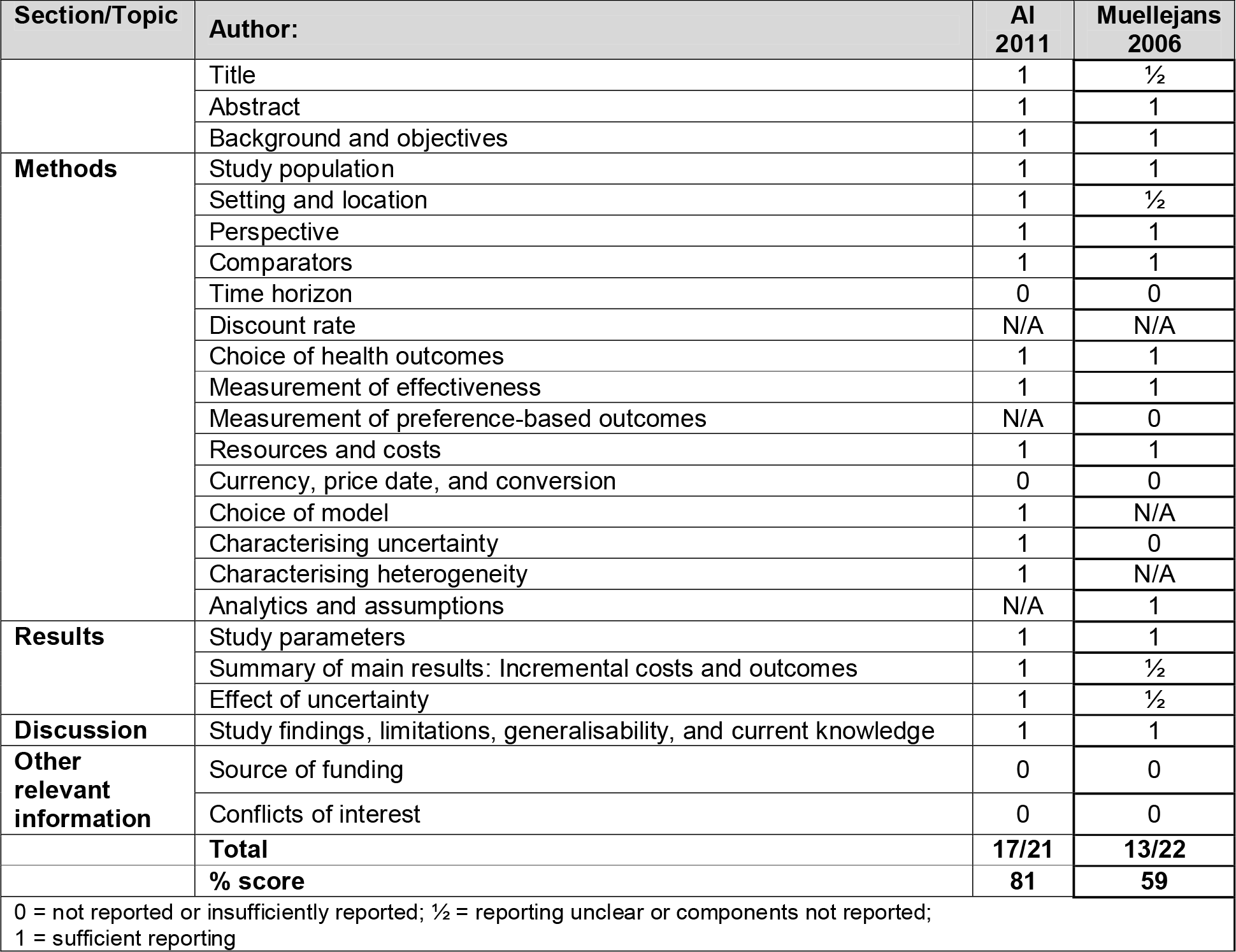
Consolidated Health Economic Evaluation Reporting Standards (CHEERS) assessment of cost-effectiveness analysis studies of remifentanil with usual care, fentanyl, or other control (n = 2).

| Section/Topic | Author: | AI 2011 | Muellejans 2006 |
| --- | --- | --- | --- |
|  | Title | 1 | ½ |
|  | Abstract | 1 | 1 |
|  | Background and objectives | 1 | 1 |
| **Methods** | Study population | 1 | 1 |
|  | Setting and location | 1 | ½ |
|  | Perspective | 1 | 1 |
|  | Comparators | 1 | 1 |
|  | Time horizon | 0 | 0 |
|  | Discount rate | N/A | N/A |
|  | Choice of health outcomes | 1 | 1 |
|  | Measurement of effectiveness | 1 | 1 |
|  | Measurement of preference-based outcomes | N/A | 0 |
|  | Resources and costs | 1 | 1 |
|  | Currency, price date, and conversion | 0 | 0 |
|  | Choice of model | 1 | N/A |
|  | Characterising uncertainty | 1 | 0 |
|  | Characterising heterogeneity | 1 | N/A |
|  | Analytics and assumptions | N/A | 1 |
| **Results** | Study parameters | 1 | 1 |
|  | Summary of main results: Incremental costs and outcomes | 1 | ½ |
|  | Effect of uncertainty | 1 | ½ |
| **Discussion** | Study findings, limitations, generalisability, and current knowledge | 1 | 1 |
| **Other relevant information** | Source of funding | 0 | 0 |
|  | Conflicts of interest | 0 | 0 |
|  | **Total** | **17/21** | **13/22** |
|  | **% score** | **81** | **59** |
| 0 = not reported or insufficiently reported; ½ = reporting unclear or components not reported;<br>1 = sufficient reporting |  |  |  |

## Discussion

This systematic review identified only two economic evaluations of remifentanil use in ICU settings. The studies varied significantly in methodology, including outcomes measured, cost components, time horizons, and perspectives. This heterogeneity precluded meta-analysis and limits the certainty of conclusions. These findings highlight important gaps in the economic evidence base for remifentanil in critical care.

Economic evidence for remifentanil use in ICU settings is limited. Many initially relevant studies were excluded due to a lack of primary cost data (i.e., were not an economic evaluation) or initiation of sedation outside the ICU (i.e., surgery or anaesthesia). Given the ICU’s unique resource demands, only studies conducted specifically within this setting were deemed suitable. Ultimately, just two studies met inclusion criteria, both comparing remifentanil to fentanyl. Each supported reduced duration of IMV and ICU stay with remifentanil use. The Dutch study^11^ reported a significant reduction in ICU costs with remifentanil, while the German study^12^ found overall costs comparable between agents. Clinical outcomes beyond length of stay were either not assessed or assumed equal. Both studies were single-centre and over a decade old, limiting their generalisability and highlighting important gaps in the current evidence base.

The scope of economic evaluations has been limited due to the scarce availability of evidence from high-quality RCTs and a lack of long-term outcomes data, including QALYs. While RCTs offer strong internal validity, they often fail to capture comprehensive resource use or extended follow-up.^21^ As a result, most economic assessments rely on modelling to estimate long-term costs and effects. Both studies included in this review^11,12^ used modelling to project outcomes and assess cost-effectiveness. Robust data are needed to better inform healthcare decision-makers about the economic impact of opioid-based sedation in ICU settings.

Despite the high costs associated with IMV and the abundance of RCTs, few cost-effectiveness analyses have compared remifentanil to other opioids in ICU settings. The reviewed studies reveal substantial gaps in the economic evaluation of opioid interventions and highlight the inherent complexity of conducting such analyses in critical care. These findings underscore the urgent need for more comprehensive assessments of both costs and outcomes.

The strengths of this systematic review included its comprehensive search strategy that covered multiple electronic databases to ensure all relevant economic evaluations of remifentanil and fentanyl had been considered. The reliability of this review was increased by the independent scrutiny of study selection, data extraction and the evaluation of reporting quality that was conducted by two separate reviewers. However, this review had several limitations. First, the CHEERS checklist assessed reporting quality but did not evaluate the appropriateness of the methods or the quality of underlying data. Second, due to methodological and reporting heterogeneity across studies, a qualitative synthesis was required in place of a quantitative meta-analysis.

## Conclusion

This systematic review identified only two studies with high heterogeneity in the economic evaluations design for remifentanil, with variation in reported results. This highlight critical gaps in research, considering the substantial economic and social impact of ICU patient care, there is a need for high-quality economic evaluations. Such evaluations are essential to enhance our comprehension of the cost-effectiveness of prevalent analgo-sedation techniques in everyday clinical settings and to guide policymakers towards efficient use of resources.

## Data Availability

All data produced in the present study are available upon reasonable request to the authors

