## Supplementary Table 1 for "Costs and cost-effectiveness of Remifentanil Versus Fentanyl for analgo-sedation in Mechanically Ventilated Adult ICU Patients"

**Supplementary Table 1:** Keyword searches in the different databases.

| **Database name** | **Platform** | **Database coverage (years)** | **Date of search** | **Total number of references found** |
| --- | --- | --- | --- | --- |
| **Embase** | **Ovid** | **1997 – 2023** | **24 October 2023** | **116** |
| #23. #8 AND #13 AND #22 ([english]/lim] AND ([adult]/lim OR [aged]/lim))  #22. #14 OR #15 OR #16 OR #16 OR #17 OR #18 OR #19 OR #20  OR #21  #21. 'IC':ab,ti.  #20. 'care, intensive':ab,ti.  #19. ((intensive OR Critical) NEAR3 unit*);ab,ti  #18. 'care critical':ab,ti.  #17. 'ICU':ab,ti.  #16. 'intensive care unit*':ab,ti.  #15. 'critical care':ab,ti.  #14. 'critical illiness':ab,ti.  #13. #9 OR #10 OR #11 OR #12  #12. 'intensive care'/:exp.  #11. 'IMV': ab,ti  #10. 'ventilation*, mechanical':ab,ti  #9. 'mechanical ventilation': ab,ti  #8. #1 OR #2 OR #3 OR #4 OR #5 OR #6 OR #7  #7. 'Fentanyl':/exp  #6. 'Remifentanil':/exp  #5. 'analgesi*,opi*':ab,ti  #4. 'opioid':ab,ti  #3. 'opiate'/exp  #2. 'analgesi*':ab,ti  #1. 'analgesia'/exp | | | | |
| **Medline All** | **Ovid** | **1997 – 2023** | **1 November July 2023** | **51** |
| 1. (Analgesia)/exp  2. (Analgesi*).tw  3. (Analgesics, Opioid)/exp  4. (Analgesi*, Opioid).tw  5. (Opioid*).tw  6. (opiate*).tw  7. (Remifentanil).mp  8. (Fentanyl).mp  9. 1 or 2 or 3 or 4 or 5 or 6 or 7 or 8  9. (mechanical ventilation).mp.  10. (mechanical ventilation*).tw.  11. (Ventilation* mechanical).tw.  12. (IMV).tw  13. (intubation).tw  14. (critical care).exp.  15. (intensive care).mp  16. (intensive care unite).mp  17. ((intensive or caitical) adj3 unit*).tw.  18. (ICU).tw  19. (IC).tw  20. (care, critical).tw  21. (care, intensive).tw  22. 9 or 10 or 11 or 12 or 13 or 14 or 15 or 16 or 17 or 18 or 19 or 20 or 21  23. 9 and 22  24. limit 23 to (“all adult [18 plus years[” and [English]) | | | | |
| **Cochrane Central Register of Controlled Trials** | **Wiley** | **1997 – 2023** | **11 September 2023** | **2** |
| **Cochrane Central Register of Controlled Trials**  #1. MeSH descriptor: [Anaglesia] explode all trees  #2. Analgesi*:ti,ab,kw. (Word variations have been searched)  #3. MeSH descriptor: [Analgesics,Opioid] explode all trees  #4. Analgesi*,Opioid:.ti,ab,kw. (Word variations have been searched)  #5. Opioid:ti,ab,kw. (Word variations have been searched)  #6. Remifentanil:ti,ab,kw. (Word variations have been searched)  #7. Fentanyl :ti,ab,kw. (Word variations have been searched)  #8. #1 or #2 or #3 or #4 or #5 or #6 or #7  #9. mechanical ventilatio*:ti,ab,kw (Word variations have been searched)  #10. ventilatio*, mechanical: ti,ab,kw (Word variations have been searched)  #11 IMV*: ti,ab,kw (Word variations have been searched)  #12 intubation: ti,ab,kw (Word variations have been searched)  #13. MeSH descriptor:[Intensive care] explode all trees  #14. MeSH descriptor:[critical care] explode all trees  #15. MeSH descriptor:[ICU] explode all trees  #16. intensive care:ti,ab,kw. (Word variations have been searched)  #17. MeSH descriptor:[ intensive care unites] explode all trees  #18. (critical* near/5(sick* or ill*)):ti,ab,kw(Word variations have been searched)  #19. ((intensive or critical) near/3 unit*):ti,ab,kw(Word variations have been searched)  #20. ICU: ti,ab,kw (Word variations have been searched)  #21. care, critical. ti,ab,kw(Word variations have been searched)  #22. care, intensive: ti,ab,kw(Word variations have been searched)  #23. #9 or #10 or #11 or #12 or #13 or #14 or #15 or #16 or #17 or #18  #24. #8 and #23 | | | | |
| **Google Scholar** | **Grey literature search** | **2000-2023** | **4 August 2023** | **2** |
| 1# (critical care[Title/Abstract]) OR ICU[Title/Abstract]) OR intensive care[Title/Abstract]) OR ("Intensive Care Units"[Mesh]) OR "Critical Care"[Mesh] OR “ICU” [Mesh])  #2 (“mechanical ventilation” [Title/Abstract]) OR IMV [Title/Abstract]) OR ventilation[Title/Abstract] )  #3 remifentanil OR fentanyl [Title/Abstract]  #4 1# and 2# and #3 and #4 | | | | |
| **Key for symbols in the search strategy**  * = wildcard  AND/OR = operators  / = indexed term term  ab = abstract  exp = explodes the term entered and retrieves records that contain the term and any of its narrower, more specific terms.  mp = title, abstract, original title, name of substance word, subject heading word, keyword heading word, protocol supplementary concept word, rare disease supplementary concept word, unique identifier, synonyms]  kw = key word  ti = title | | | | |
